# Deep learning-based identification of genetic variants: Application to Alzheimer’s disease classification

**DOI:** 10.1101/2021.07.19.21260789

**Authors:** Taeho Jo, Kwangsik Nho, Paula Bice, Andrew J. Saykin, for the Alzheimer’s Neuroimaging Initiative

**Affiliations:** Department of Radiology and Imaging Sciences, Center for Neuroimaging, Indiana University School of Medicine, Indianapolis, IN, USA; Indiana Alzheimer’s Disease Research Center, Indiana University School of Medicine, Indianapolis, IN, USA; Indiana University Network Science Institute, Bloomington, IN, USA; Center for Computational Biology and Bioinformatics, Indiana University School of Medicine, Indianapolis, IN, USA; Department of Medical and Molecular Genetics, Indiana University School of Medicine, Indianapolis, IN, USA

**Keywords:** Deep learning, genetic variants, Alzheimer’s disease, genome–wide association studies, phenotype influence scores

## Abstract

Deep learning is a promising tool that uses nonlinear transformations to extract features from high-dimensional data. Deep learning is challenging in genome–wide association studies (GWAS) with high-dimensional genomic data. Here we propose a novel three-step approach (SWAT-CNN) for identification of genetic variants using deep learning to identify phenotype-related single nucleotide polymorphisms (SNPs) that can be applied to develop accurate disease classification models. In the first step, we divided the whole genome into non-overlapping fragments of an optimal size and then ran Convolutional Neural Network (CNN) on each fragment to select phenotype-associated fragments. In the second step, using a Sliding Window Association Test (SWAT), we ran CNN on the selected fragments to calculate phenotype influence scores (PIS) and identify phenotype-associated SNPs based on PIS. In the third step, we ran CNN on all identified SNPs to develop a classification model. We tested our approach using GWAS data from the Alzheimer’s Disease Neuroimaging Initiative (ADNI) including (N=981; cognitively normal older adults (CN) =650 and AD=331). Our approach identified the well-known *APOE* region as the most significant genetic locus for AD. Our classification model achieved an area under the curve (AUC) of 0.82, which was compatible with traditional machine learning approaches, Random Forest, and XGBoost. SWAT-CNN, a novel deep learning-based genome-wide approach, identified AD-associated SNPs and a classification model for AD and may hold promise for a range of biomedical applications.

## Introduction

Deep learning is a representative machine learning algorithm that enables nonlinear transformations to extract features of high-dimensional data ^1^, unlike traditional machine learning models that predict a linear combination of weights by assuming a linear relationship between input features and a phenotype of interest. Deep learning has been used to predict disease outputs by handling original high-dimensional medical imaging data without feature selection procedures ^2, 3^. In genetic research, deep learning frameworks have been used to investigate molecular phenotypes that predict the effects of non-coding variants^4-10^, differential gene expression ^11^, and potential transcription factor binding sites ^12^. These tools use CHIP-Seq or DNase-Seq data as training data to predict chromatin features such as transcription factor binding or DNase hypersensitivity from DNA sequences. More recently, deep learning has been employed in the capture of mutations and the analysis of gene regulations, demonstrating its potential for furthering our understanding of epigenetic regulation ^13^. Furthermore, deep learning is being used in gene therapy to design CRISPR guide RNAs using deep learning-based gene features ^14-19^.

Genome-wide association studies (GWAS) use a statistical approach by considering one single nucleotide polymorphism (SNP) at a time across the whole genome to identify population-based genetic risk variation for human diseases and traits ^20, 21^. However, deep learning has not yet been used to perform GWAS, as it is challenging due to the so-called high-dimension low-sample-size (HDLSS) problem ^22^, which is known to impact phenotype prediction using genetic variation. Feature reduction approaches have been commonly used ^23-25^ to resolve this problem, but feature reduction using high-dimensional genomic data is also challenging due to a NP-hard problem^26, 27^. Therefore, it is necessary to develop a deep learning framework to identify genetic variants using whole genome data.

Here we proposed a novel three-step deep learning-based approach to select informative SNPs and develop classification models for a phenotype of interest. In the first step, we divided the whole genome into non-overlapping fragments of an optimal size and then used deep learning algorithms to select phenotype-associated fragments containing phenotype-related SNPs. Different sized fragments and several deep learning algorithms were tested to select the optimal size for fragments and the optimal algorithm. In the second step, we ran the optimal deep learning algorithm using an overlapping Sliding Window Association Test (SWAT) within selected fragments to calculate phenotype influence scores (PIS) using SNPs and the phenotype of interest to identify informative SNPs. In the third step, we ran the optimal algorithm again on all identified informative SNPs to develop a classification model.

Alzheimer’s disease (AD), the most common form of dementia, is a neurodegenerative disorder that causes progressive deterioration of memory and cognitive function. The pathological hallmarks of the disease are toxic amyloid-β plaques and neurofibrillary tau tangles in the brain^28, 29^, with the strongest genetic risk factor being the ε4 allele of apolipoprotein E (*APOE*). *APOE* ε4 allele carriers are more prone to amyloid deposition and have a 3-to 4-fold increased risk of AD^30-32^. In addition to amyloid, tau and *APOE*, many other aging and neurodegeneration associated biological pathways are being actively investigated for their role in AD pathogenesis and for their potential as targets for therapeutic development. Examples include inflammation, cellular senescence, telomere shortening, altered neurogenesis, dysregulated lipid metabolism, altered mitochondrial function and brain energetics, and other age associated factors.^33-39^ In addition, processes related to clearance of misfolded proteins are important including autophagy, the primary mechanism that removes protein aggregates^40, 41^. Relatedly, mitophagy plays an essential role in maintaining mitochondrial homeostasis and when impaired may contribute to AD related pathophysiology ^42, 43^. As amyloid-β is clearly linked to the initiation and progression of AD, it has been targeted for drug treatment. Despite longstanding global efforts and numerous failed trials, the FDA recently granted accelerated approval of aducanumab, the first potentially disease-modifying anti-amyloid treatment ^44^. In addition, it is of fundamental importance to identify biomarkers for the detection of AD at pre-symptomatic stages to slow or prevent disease progression^45-47^. In the past few years, artificial intelligence (AI) approaches has been used to identify AD biomarkers through brain image analysis^3, 48, 49^, Cerebrospinal fluid (CSF) AD biomarkers ^50^ and plasma metabolites ^51^. Using high-throughput bioassays, AI technology has been used to repurpose known drugs to treat Alzheimer’s disease ^52, 53^. Though these AI applications are growing rapidly, only a few have reached the clinical stage.

We tested our approach using only whole genome data for AD (N=981; cognitively normal older adults (CN) =650 and AD=331). Our approach identified the known *APOE* region as the most significant genetic locus for AD. Using the identified region, we made a classification model with CNN. To determine if the algorithm is comparable to traditional machine learning algorithms, we also applied XG Boost and Random Forest. Our classification model yielded 75.2% accuracy which was generally compatible with a modest gain in accuracy of 3.8% and 9.6% relative to XG Boost and Random Forest, respectively. Our classification model yielded 75.2% accuracy over traditional machine learning methods, being 3.8% and 9.6% higher than XG Boost and Random Forest, respectively. Our novel deep learning-based approach can identify informative SNPs and develop a classification model for AD by combining nearby SNPs and testing their aggregation.

## Materials and Methods

### Study participants

All individuals used in the analysis were participants of the Alzheimer’s Disease Neuroimaging Initiative (ADNI) cohort ^54, 55^. The ADNI initial phase (ADNI-1) was launched in 2003 to test whether serial magnetic resonance imaging (MRI), position emission tomography (PET), other biological markers, and clinical and neuropsychological assessment could be combined to measure the progression of Mild Cognitive Impairment (MCI) and early AD. ADNI-1 has been extended in subsequent phases (ADNI-GO, ADNI-2, and ADNI-3) for follow-up of existing participants and additional new enrollments. Demographic information, *APOE* and whole-genome genotyping data, and clinical information are publicly available from the ADNI data repository (www.loni.usc.edu/ADNI/). Informed consent was obtained for all subjects, and the study was approved by the relevant institutional review board at each data acquisition site.

### Genotyping and imputation

ADNI participants were genotyped using several Illumina genotyping platforms including Illumina Human610-Quad BeadChip, Illumina HumanOmni Express BeadChip, and Illumina HumanOmni 2.5M BeadChip ^56^. As ADNI used different genotyping platforms, we performed quality control procedures (QC) on each genotyping platform data separately and then imputed un-genotyped single nucleotide polymorphisms (SNPs) separately using MACH and the Haplotype Reference Consortium (HRC) data as a reference panel ^57^. Before imputation, we performed QC for samples and SNPs as described previously: (1) for SNP, SNP call rate < 95%, Hardy-Weinberg *P* value < 1×10^−6^, and minor allele frequency (MAF) < 1%; (2) for sample, sex inconsistencies, and sample call rate < 95% ^58^. Furthermore, in order to prevent spurious associations due to population stratification, we selected only non-Hispanic participants of European ancestry that clustered with HapMap CEU (Utah residents with Northern and Western European ancestry from the CEPH collection) or TSI (Toscani in Italia) populations using multidimensional scaling (MDS) analysis and the HapMap genotype data ^58, 59^. After imputation, we performed standard QC on imputed genotype data as described previously ^60^. Specifically, we imposed an r^2^ value equal to 0.30 as the threshold to accept the imputed genotypes. In the study, imputed genome-wide genotyping data from 981 ADNI non-Hispanic participants (650 cognitive normal older adults (CN) and 331 AD patients) were used with a total of 5,398,183 SNPs (minor allele frequency (MAF) > 5%).

### Genome-wide association study (GWAS)

Using imputed genotypes, a GWAS for AD was conducted. For the GWAS, logistic regression with age and sex as covariates was performed using PLINK^61^ to determine the association of each SNP with AD. To adjust for multiple testing, a conservative threshold for genome-wide significant association (*p* < 5 × 10^−8^) was employed based on a Bonferroni correction.

### Fragmentation of whole genome data

Whole genome data for 981 participants were divided into non-overlapping fragments of varying sizes from 10 SNPs to 200 SNPs to determine the optimal fragmentation size. The sub-data sets consisting of fragments of the same size were divided into train-test-validation sets (60:20:20), and Convolutional Neural Network (CNN)^62^, Long short-term memory (LSTM)^63^, LSTM-CNN^64^, and Attention^65^ algorithms were applied to each. Early stopping using a validation set was applied to prevent over-fitting, followed by the measurement of training time and accuracy (ACC).

### Deep learning on fragments

Deep learning is the result of continuous development such as perceptron^66, 67^, which adds the concept of weight adjustment to the theory that it can behave like a human brain when neurons with on-off functions are connected in a network form^68^, and Adaline^69^, which uses gradient descent to update weights. These early neural nets were advanced to a multilayer perceptron, which includes hidden layers to solve the famous XOR problem^70^, marking a theoretical turning point with the concept of backpropagation to update the weight of the hidden layer^71-74^. The inherent problem of backpropagation, in which vanishing gradients occur when there are many layers^75^, has been alleviated through activation functions, such as sigmoid function and ReLU^76, 77^, as well as optimization methods for better gradient descent methods, such as Ada-Grad^78^, RMSprop^79^, and Adam^80^. These developments, along with the advancement of GPU hardware, have created an era of deep learning as it is now.

Deep learning has laid the theoretical foundation for backpropagation, the application of activation functions, and the development of optimizers for better gradient descent. Common deep learning algorithms, such as CNN, LSTM, and Attention, have a hierarchical structure that implements an enhanced version of the basic principles of deep learning. The detailed technical description of each algorithm is described extensively in the relevant paper, so here we focus on the core of the deep learning technology commonly applied to the algorithm used in the experiment.

We used ReLU as an activation function that underlies the deep learning algorithms used in our experiments. ReLU, the most used activation function in the deep learning community, replaces the given value with zero if the value is < 0 and uses the given value if it is > 0. Thus, if the given value is greater than zero, the derivative becomes one, and the weight can be adjusted without vanishing the gradient to the first layer through the hidden layer. We used Adam as the optimization method. Adam is currently the most popular optimization method for deep learning, as it takes advantage of momentum SGD^81^ and RMSprop, which are expressed as follows: G_t_is the sum of the square of the modified gradient, and ε is a very small constant that prevents the equation from being divided by zero.

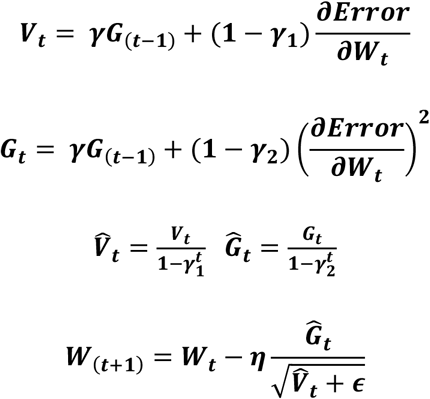

Backpropagation is used to calculate the initial error value from a given random weight using the least squares method and then to update the weight using a chain rule until the differential value becomes zero. Here, the differential value of zero means that the weight does not change when the gradient is subtracted from the previous weight.

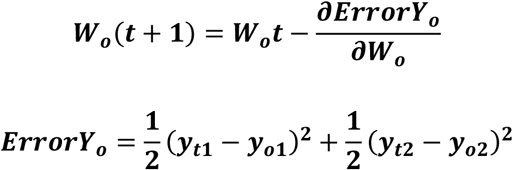

If y_o1_and y_o2_are the output values of the output layer coming through the hidden layer and the actual values of the given data are y_t1_and y_t2_, the partial derivative of the error ErrorY_o_to the weight of the output layer can be calculated using the chain rule as follows:

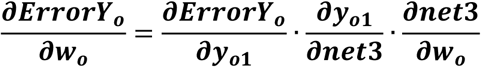

The partial derivative of the error ErrorY_o_to the weight of the hidden layer can be calculated as follows:

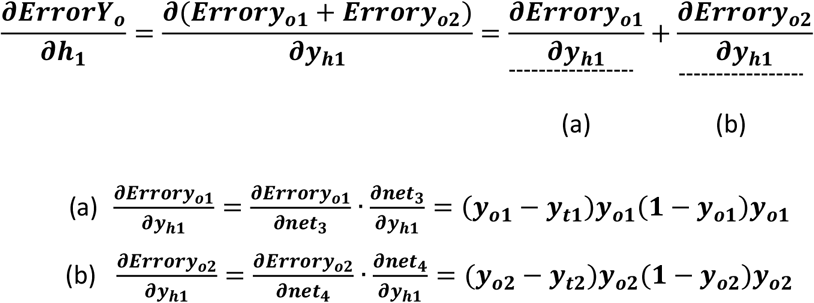

Accordingly, the weight w_h_of the hidden layer is updated as follows:

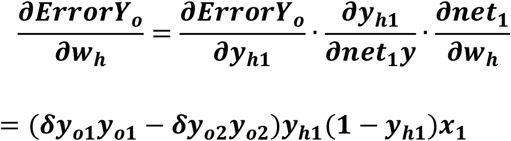

### Calculation of phenotype influence score using deep learning

Prediction accuracy was calculated from deep learning applied to each fragment and converted to a z-score. The z-score follows a normal distribution with µ = 1 and σ = 0, under the hypothesis that there is no relationship between the variables in the population. Fragments with a z-score higher than the median were selected. An overlapping SWAT for the calculation of PIS is applied to these fragments. When the length of the fragment is w, the window is positioned w-1 from the first SNP of the fragment and moves by one SNP and stops at the last SNP of the fragment. Each region within the SWAT is divided into a train-test-validation set (60:20:20), and early stopping using a validation set is applied to prevent over-fitting. When the kth SNP is S_k_, PIS is calculated as follows.

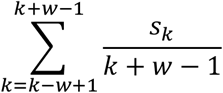

This SWAT is applied to all selected fragments, resulting in a PIS score for all SNPs.

### Phenotype classification using deep learning

We selected the top 100 to 10,000 SNPs based on the PIS. For the AD-CN classification, we used a CNN consisting of convolution layers with a kernel size of 5, pooling the layer with a max-pool size of 2, a fully connected layer of 64 nodes, and an output layer with a softmax activation function. Due to gradient vanishing and explosion issues caused by the repeated multiplication of the recurrent weight matrix, RNN or its variants had difficulty training. In order to compare the performance, we also applied Random Forest and XG Boost, which are traditionally used for tabular data classification. XG Boost is a tree-based ensemble algorithm, one of popular implementations of gradient boosting. We trained XGboost using a “xgboost” package for python (https://xgboost.readthedocs.io/). Random Forest is another ensemble learning method that uses many decision trees as its classifiers^82, 83^. We trained Random Forest using the scikit-learn package for python by setting the number of trees as 10 and the maximum depth of each tree as 3.

## Results

Our deep learning-based approach consists of three steps to select informative SNPs and develop an accurate classification model. In the first step, we divided the whole genome into non-overlapping fragments of an optimal size. To choose an optimal fragment size and an optimal deep learning algorithm, we calculated the mean accuracy and computation time for classification of AD using various fragment sizes containing 10 to 200 SNPs and several deep learning algorithms (CNN, LSTM, LSTM-CNN, Attention). In this analysis, we used 10-200 SNPs located within a region surrounding the *APOE* gene, the strongest and most robust AD genetic risk locus. Figure 1 shows the average accuracy and computation time for CNN, LSTM, LSTM-CNN, and Attention as a function of the fragment size. As shown in Fig. 1A, the analysis yielded the highest accuracy for classification of AD for a fragment size with 40 SNPs (Fig. 1A). Fig 1B shows the average accuracy and time as a function of the deep learning algorithm on window size of 40 within a region surrounding the *APOE* gene. CNN and LSTM-CNN models had the highest accuracy for classification of AD, followed by LSTM. However, the computation time of CNN and LSTM models were 5.9 seconds and 40.4 seconds, respectively. The computation time of LSTM, LSTM-CNN, and Attention models sharply increased compared to CNN because the fragment contains more SNPs; therefore, we chose a fragment with 40 SNPs as the optimal fragment size for the CNN and optimal deep learning algorithm, respectively. The whole genome was divided into 134,955 fragments, each with 40 SNPs. We ran CNN on each fragment to calculate z-scores based on classification accuracy and selected phenotype-associated fragments. We selected 1,802 fragments with z-scores higher than a median z-score.

**[Figure 1].**
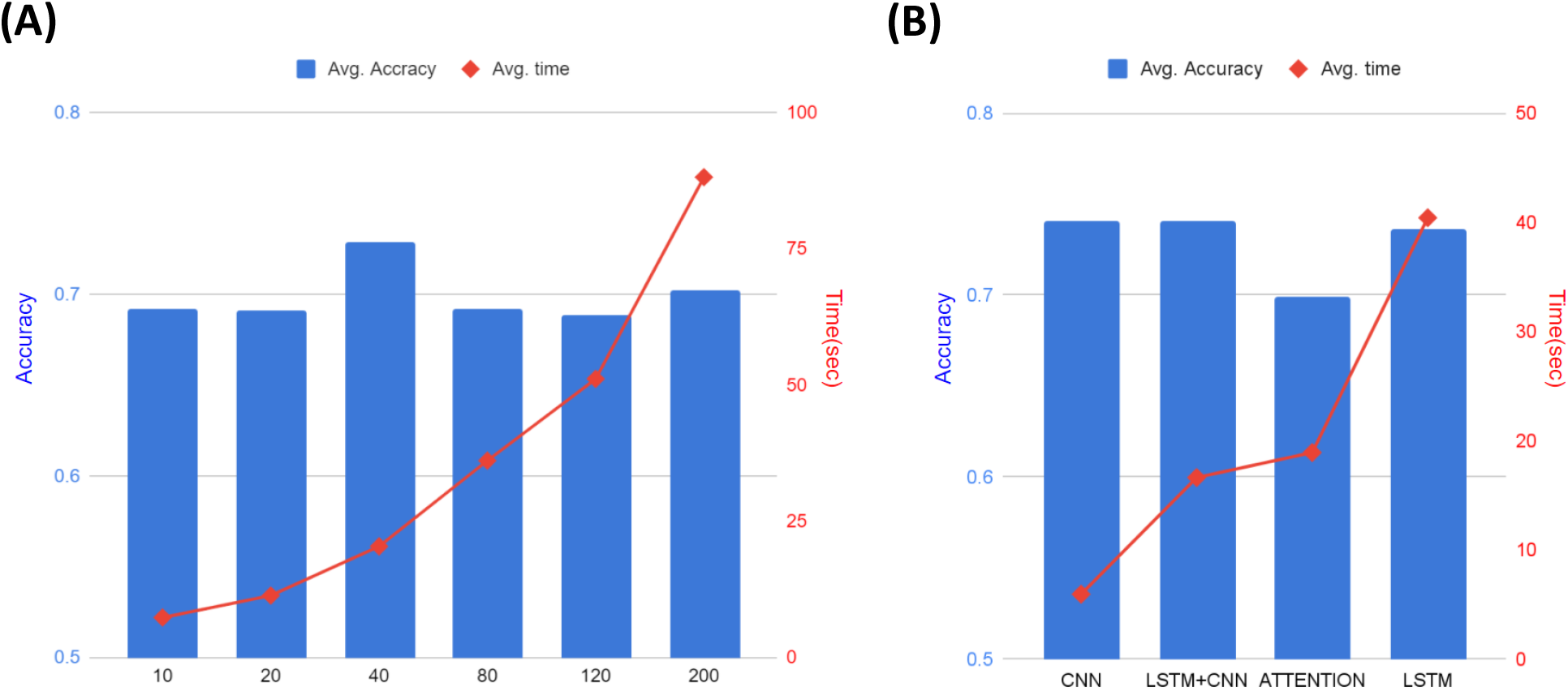
Selection of an optimal fragment size and an optimal deep learning algorithm. In order to choose an optimal fragment size and an optimal deep learning algorithm, the mean accuracy and computation time was calculated for the classification of AD using various fragment sizes containing 10 to 200 SNPs in the *APOE* region and several deep learning algorithms (CNN, LSTM, LSTM-CNN, and Attention). (A) Average accuracy and time as a function of the fragment size. The highest accuracy for classification of AD was obtained with a fragment having 40 SNPs in CNN, LSTM-CNN and LSTM models. The accuracy difference was not large according to window size, but the processing time increased with window size. (B) Average accuracy and time as a function of the deep learning algorithm using a widow size of 40. The computation time of LSTM, LSTM-CNN and Attention models increases sharply compared to CNN as they include more SNPs in their fragments.

In the second step, using a SWAT, we ran CNN on the selected fragments to calculate the PIS of each SNP in the selected fragments and identify phenotype-associated SNPs based on the PIS, as shown in Fig. 2. For each SNP, we calculated a mean accuracy of 40 windows, which is the PIS of the SNP. Using PIS values, we calculated the z-scores and one-tailed p-values. Figure 3 shows a Manhattan plot with the -log10 p-values on the y-axis against the SNP position in the genome on the x-axis. The SNP with the smallest p-value was rs5117 in the *APOC1* gene (p-value=1.04E-22) and rs429358 in the *APOE* gene (p-value of 1.41E-16). The genetic region including *APOE*/*APOC1*/*TOMM40* genes is known as the strongest genetic risk locus for AD^30, 84-86^. The next highest genetic loci were located at *SNX14, SNX16, BICD1, WDR72*, and *GLT1D1* genes.

**[Figure 2].**
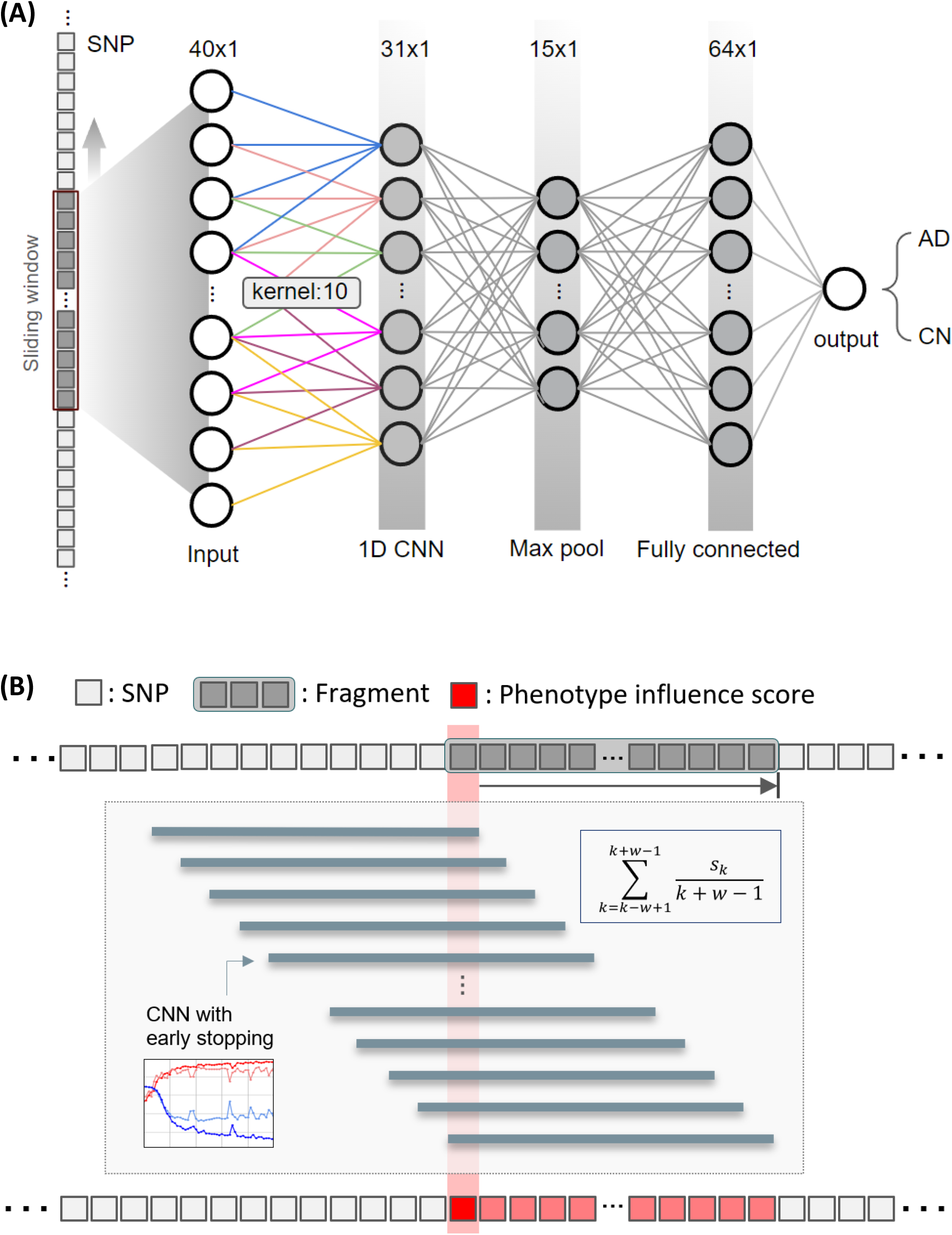
Sliding Window Association Test (SWAT) for genetic variants. (A) Inside view of a sliding window that traverses the entire genome sequence to find a location that is associated with a specific phenotype. A CNN consisting of a convolutional layer with a kernel size of 10, a pooling layer with a maximum pool size of 2, a fully connected layer of 64 nodes, and an output layer with softmax activation was used. (B) Framework to calculate phenotype influence scores of SNPs. We divided the whole genome into 134,955 fragments, each with 40 SNPs. To calculate a phenotype influence score for each of the 40 SNPs included in one fragment, we used an overlapping window approach and CNN. w is the number of SNPs in the fragment and S_k_is the k_th_SNP in the fragment.

**[Figure 3].**
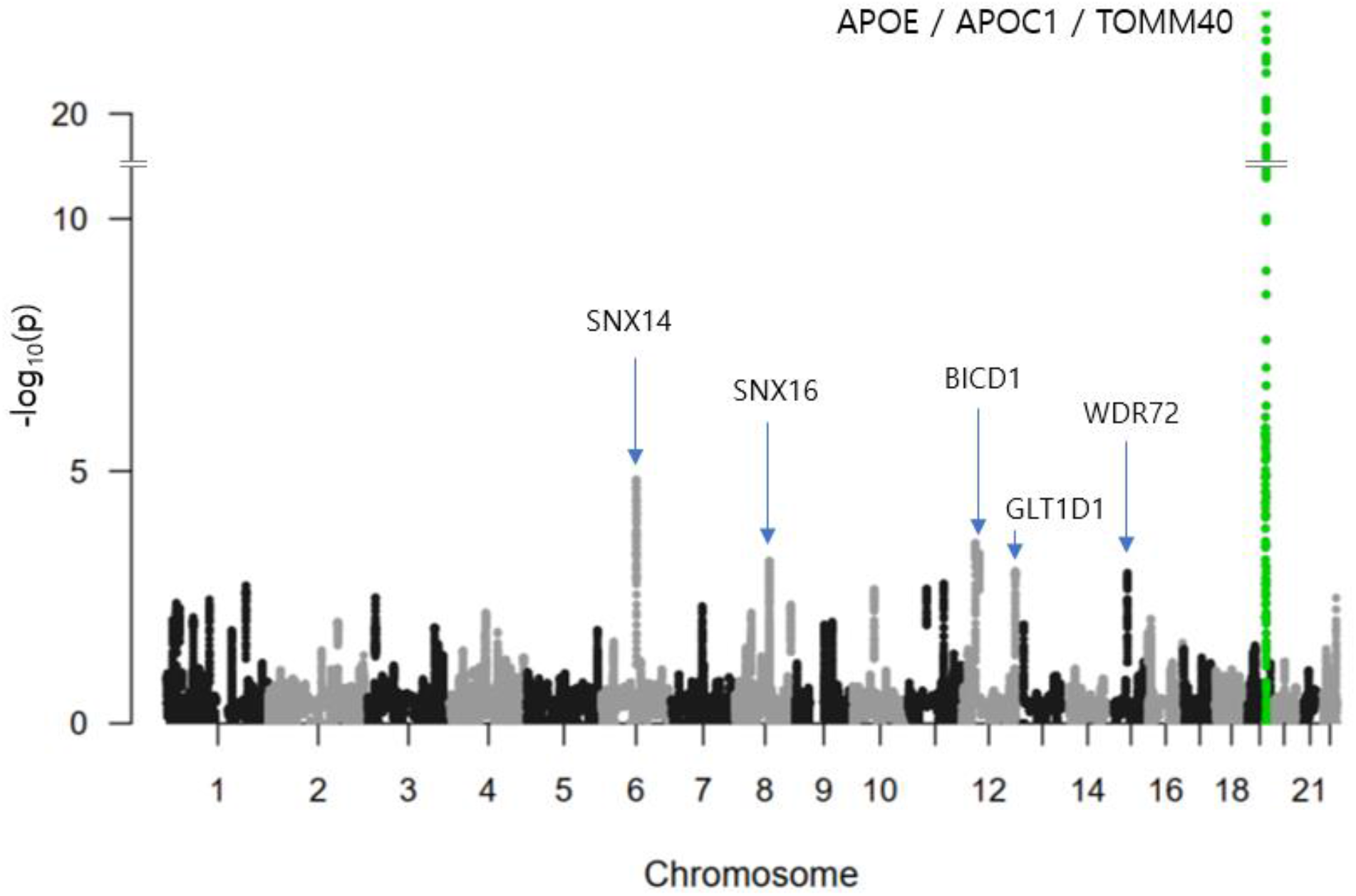
Manhattan plot of p-values of SNPs by our deep learning-based approach in AD. The X-axis shows SNP positions in the genome. The Y-axis shows -log10 of p-values. The genetic region including *APOE, APOC1*, and *TOMM40* genes is known as the strongest genetic risk locus for Alzheimer’s disease. The SNP with the smallest p-value was rs5117 in *APOC1* gene (P=1.04E-22). rs429358 in *APOE* has a p-value of 1.41E-16. Next identified genetic loci were located at *SNX14, SNX16, BICD1, WDR72*, and *GLT1D1* genes.

In the third step, we ran CNN on the identified SNPs to develop an AD classification model. Table 1 shows the classification results of AD vs. CN using subsets containing the top 100 to 10,000 SNPs based on PIS. For comparison with traditional machine learning methods, we used two popular algorithms as classifiers, XG Boost and Random Forest. The highest mean accuracy of 10-cross validation in classifying AD from CN by CNN was 75.02% (area under the curve [AUC] of 0.8157) for a subset containing 4,000 SNPs, which had a 6.3% higher accuracy than Random Forest for a subset containing 2,000 SNPs and a 1.94% higher accuracy than XG Boost for a subset containing 1,000 SNPs. When we calculated the classification accuracy of AD using only the number of *APOE* ε4 alleles, the classification accuracy was 66.7%, which was 8.3% lower than our method. Our CNN models outperformed two traditional machine learning models, Random Forest and XGBoost, in all cases as shown in Fig. 4.

**[Table 1].** Results of classification of AD from CN. The table shows the number of top SNPs selected based on phenotype influence score for AD classification and the accuracy and AUC of 10-fold cross-validation. Our CNN-based approach yielded the highest accuracy and AUC of 75.02% and 0.8157, respectively, for 4,000 SNPs. In all cases, our CNN models outperformed two traditional machine learning models, Random Forest and XG Boost.

| Top | Random Forest |  |  |  | XG Boost |  |  |  | CNN |  |  |  |
| --- | --- | --- | --- | --- | --- | --- | --- | --- | --- | --- | --- | --- |
| | Accuracy | STD(±) | AUC | STD(±) | Accuracy | STD(±) | AUC | STD(±) | Accuracy | STD(±) | AUC | STD(±) |
| **100** | 66.46 | 7.79 | 71.37 | 5.76 | 70.24 | 2.80 | 72.66 | 2.81 | 68.29 | 2.87 | 72.16 | 6.03 |
| **200** | 67.18 | 3.88 | 71.75 | 3.86 | 67.99 | 1.52 | 71.66 | 2.45 | 69.52 | 5.08 | 71.82 | 4.94 |
| **300** | 66.26 | 3.80 | 70.98 | 3.77 | 68.20 | 3.32 | 70.29 | 2.72 | 70.64 | 2.20 | 72.50 | 5.85 |
| **400** | 67.58 | 4.67 | 70.74 | 4.28 | 69.42 | 3.43 | 71.77 | 2.34 | 67.99 | 4.65 | 71.67 | 4.12 |
| **500** | 67.59 | 7.79 | 71.11 | 4.57 | 71.05 | 2.56 | 73.81 | 3.25 | 71.56 | 6.58 | 74.11 | 6.14 |
| **1000** | 68.31 | 5.22 | 71.78 | 4.45 | **73.08** | 2.89 | 74.07 | 3.72 | 73.91 | 3.87 | 77.41 | 4.44 |
| **2000** | **68.70** | 3.13 | **73.72** | 4.24 | 72.48 | 2.61 | **75.09** | 3.65 | 73.29 | 2.77 | 77.82 | 4.09 |
| **3000** | 67.78 | 3.59 | 72.82 | 3.51 | 69.62 | 4.27 | 73.76 | 3.28 | 73.80 | 2.40 | 78.62 | 2.82 |
| **4000** | 68.19 | 4.69 | 72.63 | 4.69 | 71.15 | 4.07 | 74.12 | 3.68 | **75.02** | 3.17 | **81.57** | 2.61 |
| **5000** | 66.25 | 5.41 | 71.05 | 3.99 | 70.74 | 3.14 | 73.30 | 3.05 | 73.19 | 4.72 | 80.03 | 5.06 |
| **10000** | 66.26 | 5.59 | 69.19 | 5.28 | 69.63 | 3.27 | 72.48 | 2.11 | 71.05 | 6.57 | 70.83 | 14.24 |

**[Figure 4].**
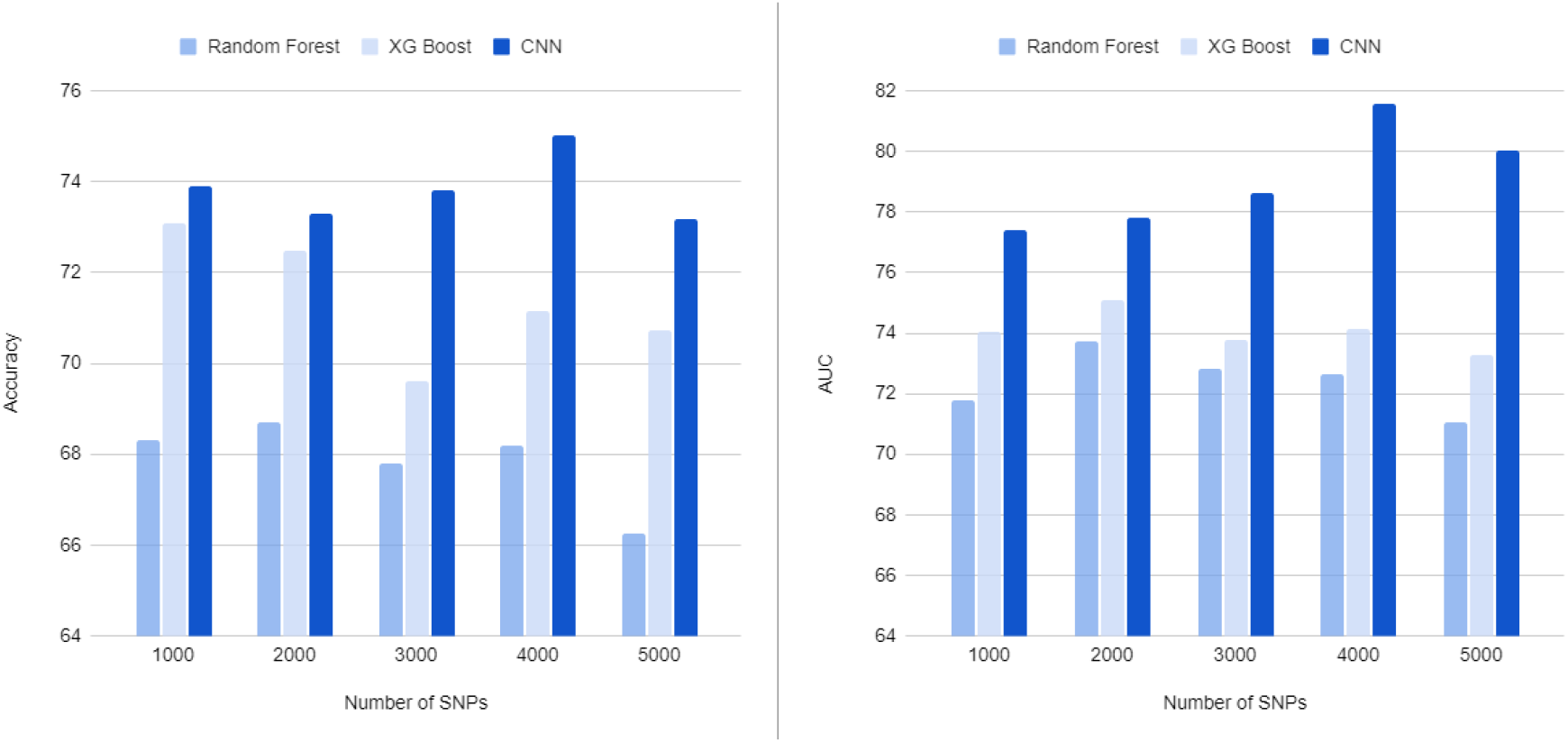
Results of classification of AD from CN. The X-axis shows the number of top SNPs selected based on phenotype influence score for AD classification. The Y-axis shows the accuracy (A) and AUC (B) of 10-fold cross-validation. Our CNN-based approach yielded the highest accuracy and AUC of 75.02% and 0.8157, respectively, for 4,000 SNPs. In all cases, our CNN models outperformed two traditional machine learning models, Random Forest and XG Boost.

Figure 5 shows LocusZoom plots^87^ for SNPs located at 300 kb upstream and downstream regions from the boundary of the *APOE* gene. The horizontal axis is the location of SNPs, and the vertical axis is -log10 of the p-values. Each dot represents a SNP, and the color represents the squared correlation coefficient (r^2^) with the most significant SNP. Figure 5A shows p-values calculated using PLINK, and the most significant SNP was rs429358 in *APOE*. Figure 5B shows p-values calculated using our deep learning approach, and the most significant SNP was rs5117 in *APOC1*. Fig. 5B shows a linear increase on the left side of rs5117 and a linear decrease on the right side of rs5117, which was different from PLINK results (Fig. 5A), which has no linear patterns. In addition, Fig. 5B, shows three strongly correlated SNPs (r^2^>0.8), with rs5117 on the left side of rs5117 but no SNPs on the right side of rs5117.

**[Figure 5].**
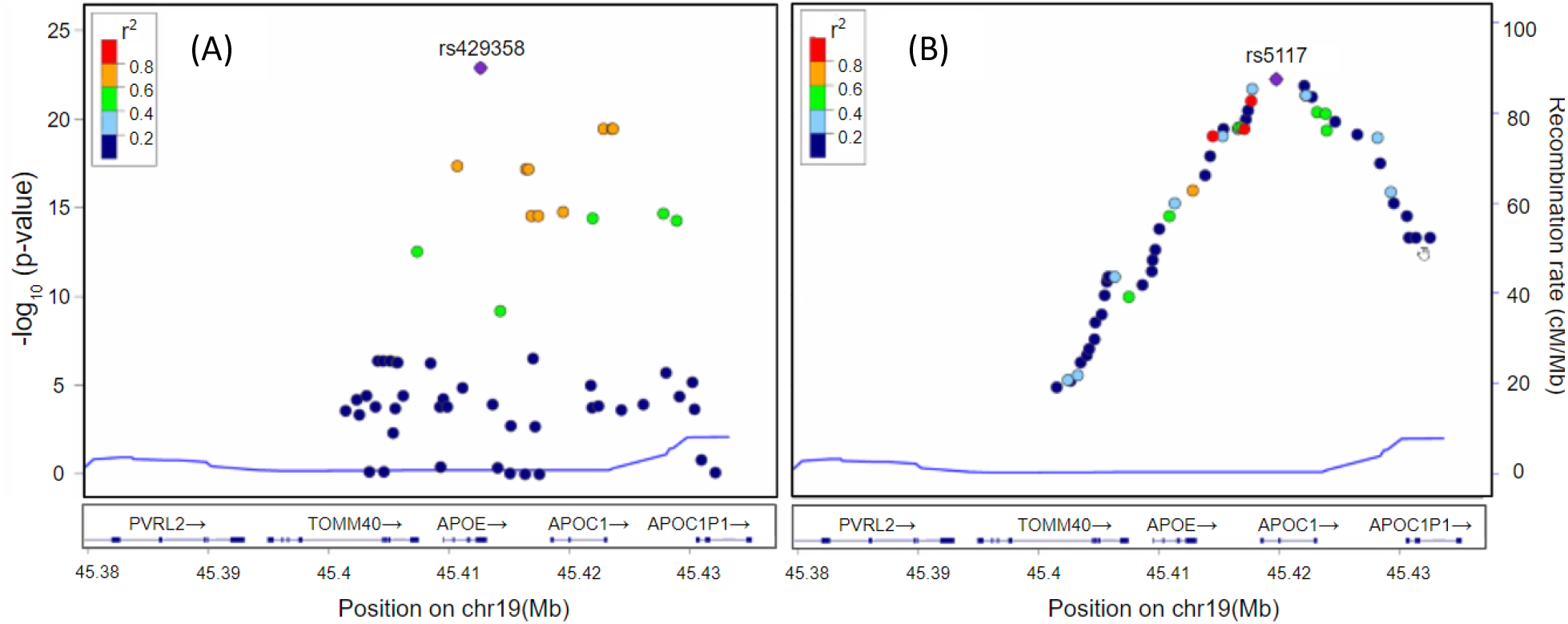
LocusZoom plots for SNPs located at the 300 kb upstream and downstream region from the boundary *APOE* gene. The horizontal axis is the location of SNPs and the vertical axis is -log10 of p-values. Each dot represents a SNP and the color represents the squared correlation coefficient (r^2^) with the most significant SNP. (A) shows p-values calculated using PLINK and the most significant SNP was rs429358 in *APOE*. (B) shows p-values calculated using our deep learning approach and the most significant SNP was rs5117 in *APOC1*. In (B), there is a linear increase on the left side of rs5117 and a linear decrease on the right side of rs5117 that was different from the PLINK results (A), which have no linear patterns. In addition, in (B), there are three strongly correlated SNPs (r^2^>0.8) with rs5117 on the left side of rs5117 but no SNPs on the right side of rs5117.

## Discussion

Although deep learning has solved many real-world problems, few deep learning approaches have been used in GWAS or sequence data to identify genetic variants and for disease/risk classification due to the high dimensionality of the genomic data ^22^. In this study, we propose a novel deep learning-based sliding window approach to identify and select disease-associated SNPs and develop an accurate classification model using high dimensional genomic data that we tested using the ADNI cohort (N=981). The proposed method successfully identified significant genetic loci for AD that included the well-known *APOE* genetic locus and highlighted several other risk loci. Our deep learning-based approach was compatible to traditional machine learning methods for classification of AD.

The deep learning-based approach for identification of genetic variants consists of three steps. In the first step, we divided the whole genome into non-overlapping fragments of an optimal size, creating a fragmentation and windowing approach that, to the best of our knowledge, is the first deep learning-based method for identifying genetic variants.

In the second step, we calculated a PIS of each SNP within the selected fragments by using an overlapping window and CNN algorithm. Our method calculates PIS, a novel index which is used to find disease-related variants and predict disease. Furthermore, we calculated the z-scores and one-tailed p-values using PIS, which yielded a Manhattan plot showing the most significant genetic loci in *APOE*/*APOC1*/*TOMM40* genes that are known as the strongest genetic risk factors for AD. Our method also identified several novel candidate genetic loci, including sorting nexin (SNX) *14* and *SNX16, located* on chromosomes 6 and 8, respectively, that have not been previously identified to be associated with AD, though there may be special relevance for neurodegeneration as *SNX12*^*88*^, *SNX17*^*89*^, *SNX27*^90^, and *SNX33*^*91*^ are involved in neuronal survival. Bicaudal D1 (*BICD1*) on chromosome 12 is a susceptibility gene in chronic obstructive pulmonary disease^92^ and lissencephaly^93^, but there are no reports of it being associated with AD.

In the third step, we selected top SNPs based on PIS to develop classification models for AD. We selected sets of highly AD-related SNPs and classified AD from CN using CNN, as well as two popular traditional machine learning algorithms, XGBoost and Random Forest. We found the accuracy of classification was changed with the number of the selected SNPs and the classification algorithms. The highest mean accuracy of the classification was 75.0% when CNN was used on the top 4,000 SNPs, which was comparable to two traditional machine learning algorithms. It was also 8.3% higher than the accuracy of the classification using only the number of *APOE* ε4 alleles. Classification is the first step toward achieving a better understanding of the genetic architecture of AD. The proposed method will benefit from future studies that use deep learning with quantitative phenotypes and baseline values to predict future disease trajectories.

We plotted the SNPs selected by PIS and PLINK for comparison using LocusZoom. We found that there were no SNPs with r^2^ greater than 0.8 in the PLINK results, but three strongly associated SNPs were identified using our method. This is because the PLINK method finds statistical significance SNP by SNP, whereas the deep learning approach uses multiple inputs to adjust weights through the training process. Deep learning uses adjacent SNPs to compute gradients at every epoch and uses a loss function to adjust the weights in the backpropagation process. Unlike PLINK, our method shows that SNPS related to a phenotype can be extracted by considering surrounding SNPs, which means that both methods might be complementary because they identify different variants, though notably in the same region around *APOE*.

In summary, our novel deep learning-based approach can identify AD-related SNPs by using genome-wide data to develop a classification model for AD. The heritability of AD is estimated to be up to 80%. Accordingly, it is important to identify novel genetic loci related to the disease. Using a modest sample size, we found a significant genetic locus and a classification accuracy of 75%. In future work, we plan to apply our method to large-scale whole genome sequencing data sets that are expected to become available soon to identify novel AD-related SNPs and develop more accurate classification models. We also plan to study early stages of disease including mild cognitive impairment and subjective cognitive decline, where there is considerable heterogeneity and more refined classification of risk for progression to AD would be valuable. In addition, future studies will investigate use of quantitative endophenotypes that may be more informative than binary classification given their potential to elucidate genetic risk related to specific disease pathways and mechanisms.

## Key Points

- Although deep learning has been successfully applied to many scientific fields, deep learning has not been used in genome–wide association studies (GWAS) in practice due to the high dimensionality of genomic data.
- To overcome this challenge, we propose a novel three-step approach (SWAT-CNN) for identification of genetic variants using deep learning to identify phenotype-related single nucleotide polymorphisms (SNPs) that can be applied to develop accurate disease classification models.
- To accomplish this, we divided the whole genome into non-overlapping fragments of an optimal size and ran a deep learning algorithm on each fragment to select disease-associated fragments.
- We calculated phenotype influence scores (PIS) of each SNP within selected fragments to identify disease-associated significant SNPs and developed a disease classification model by using overlapping window and deep learning algorithms.
- In the application of our method to Alzheimer’s disease (AD), we identified well-known significant genetic loci for AD and achieved higher classification accuracies than traditional machine learning methods.

## Data Availability

Data from the ADNI are available from the ADNI website (adni.loni.usc.edu).

## Acknowledgements

Data collection and sharing for this project was funded by the Alzheimer’s Disease Neuroimaging Initiative (ADNI) (National Institutes of Health Grant U01AG024904) and DODADNI (Department of Defense award number W81XWH-12-2-0012). ADNI is funded by the National Institute on Aging, the National Institute of Biomedical Imaging and Bioengineering, and through generous contributions from the following: AbbVie, Alzheimer’s Association; Alzheimer’s Drug Discovery Foundation; Araclon Biotech; BioClinica, Inc.; Biogen; Bristol-Myers Squibb Company; CereSpir, Inc.; Cogstate; Eisai Inc.; Elan Pharmaceuticals, Inc.; Eli Lilly and Company; EuroImmun; F. Hoffmann-La Roche Ltd and its affiliated company Genentech, Inc.; Fujirebio; GE Healthcare; IXICO Ltd.; Janssen Alzheimer Immunotherapy Research & Development, LLC.; Johnson & Johnson Pharmaceutical Research & Development LLC.; Lumosity; Lundbeck; Merck & Co., Inc.; Meso Scale Diagnostics, LLC.; NeuroRx Research; Neurotrack Technologies; Novartis Pharmaceuticals Corporation; Pfizer Inc.; Piramal Imaging; Servier; Takeda Pharmaceutical Company; and Transition Therapeutics. The Canadian Institutes of Health Research is providing funds to support ADNI clinical sites in Canada. Private sector contributions are facilitated by the Foundation for the National Institutes of Health (http://www.fnih.org). The grantee organization is the Northern California Institute for Research and Education, and the study is coordinated by the Alzheimer’s Therapeutic Research Institute at the University of Southern California. ADNI data are disseminated by the Laboratory for Neuro Imaging at the University of Southern California.

## Author Contributions

TJ, KN, and AS: Conceptualization. AS: Acquisition of Data and Interpretation of Results. TJ, KN: Data Curation. TJ: Formal Analysis, Investigation, Methodology, Validation, Visualization. TJ, KN, PB and AS: Writing -Original Draft Preparation. TJ, KN, PB and AS: Review & Editing.

## REFERENCE

1. Wainberg M, Merico D, Delong A, Frey BJ. Deep learning in biomedicine. Nature Biotechnology 2018;36:829–838.

2. Jo T, Nho K, Saykin AJ. Deep learning in Alzheimer’s disease: diagnostic classification and prognostic prediction using neuroimaging data. Frontiers in aging neuroscience 2019;11:220.

3. Jo T, Nho K, Risacher SL, Saykin AJ, for the Alzheimer’s Neuroimaging I. Deep learning detection of informative features in tau PET for Alzheimer’s disease classification. BMC Bioinformatics 2020;21:496.

4. Zhang Z, Park CY, Theesfeld CL, Troyanskaya OG. An automated framework for efficiently designing deep convolutional neural networks in genomics. Nature Machine Intelligence 2021.

5. Zhou J, Troyanskaya OG. Predicting effects of noncoding variants with deep learning–based sequence model. Nature Methods 2015;12:931–934.

6. Alipanahi B, Delong A, Weirauch MT, Frey BJ. Predicting the sequence specificities of DNA-and RNA-binding proteins by deep learning. Nature Biotechnology 2015;33:831–838.

7. Xiong HY, Alipanahi B, Lee LJ, et al. The human splicing code reveals new insights into the genetic determinants of disease. Science 2015;347:1254806.

8. Quang D, Xie X. DanQ: a hybrid convolutional and recurrent deep neural network for quantifying the function of DNA sequences. Nucleic Acids Research 2016;44:e107–e107.

9. Angermueller C, Lee HJ, Reik W, Stegle O. DeepCpG: accurate prediction of single-cell DNA methylation states using deep learning. Genome Biology 2017;18:67.

10. Zhang S, Hu H, Jiang T, Zhang L, Zeng J. TITER: predicting translation initiation sites by deep learning. Bioinformatics 2017;33:i234–i242.

11. Tasaki S, Gaiteri C, Mostafavi S, Wang Y. Deep learning decodes the principles of differential gene expression. Nature Machine Intelligence 2020;2:376–386.

12. Zheng A, Lamkin M, Zhao H, Wu C, Su H, Gymrek M. Deep neural networks identify sequence context features predictive of transcription factor binding. Nature Machine Intelligence 2021;3:172–180.

13. Scherer M, Schmidt F, Lazareva O, et al. Machine learning for deciphering cell heterogeneity and gene regulation. Nature Computational Science 2021;1:183–191.

14. Listgarten J, Weinstein M, Kleinstiver BP, et al. Prediction of off-target activities for the end-to-end design of CRISPR guide RNAs. Nature Biomedical Engineering 2018;2:38–47.

15. Shen MW, Arbab M, Hsu JY, et al. Predictable and precise template-free CRISPR editing of pathogenic variants. Nature 2018;563:646–651.

16. Leenay RT, Aghazadeh A, Hiatt J, et al. Large dataset enables prediction of repair after CRISPR– Cas9 editing in primary T cells. Nature Biotechnology 2019;37:1034–1037.

17. Liu Q, He D, Xie L. Prediction of off-target specificity and cell-specific fitness of CRISPR-Cas System using attention boosted deep learning and network-based gene feature. PLOS Computational Biology 2019;15:e1007480.

18. Kim HK, Min S, Song M, et al. Deep learning improves prediction of CRISPR–Cpf1 guide RNA activity. Nature Biotechnology 2018;36:239–241.

19. Ogden PJ, Kelsic ED, Sinai S, Church GM. Comprehensive AAV capsid fitness landscape reveals a viral gene and enables machine-guided design. Science 2019;366:1139.

20. Yan J, Qiu Y, Ribeiro dos Santos AM, et al. Systematic analysis of binding of transcription factors to noncoding variants. Nature 2021;591:147–151.

21. Buniello A, MacArthur JA L, Cerezo M, et al. The NHGRI-EBI GWAS Catalog of published genome-wide association studies, targeted arrays and summary statistics 2019. Nucleic Acids Research 2019;47:D1005–D1012.

22. Auton A, Abecasis GR, Altshuler DM, et al. A global reference for human genetic variation. Nature 2015;526:68–74.

23. Li F, Yang Y, Xing EP. From Lasso regression to feature vector machine. Proceedings of the 18th International Conference on Neural Information Processing Systems. Vancouver, British Columbia, Canada: MIT Press, 2005: 779–786.

24. Yamada M, Jitkrittum W, Sigal L, Xing EP, Sugiyama M. High-Dimensional Feature Selection by Feature-Wise Kernelized Lasso. Neural Computation 2014;26:185–207.

25. Xu Z, Huang G, Weinberger KQ, Zheng AX. Gradient boosted feature selection. Proceedings of the 20th ACM SIGKDD international conference on Knowledge discovery and data mining. New York, New York, USA: Association for Computing Machinery, 2014: 522–531.

26. Amaldi E, Kann V. On the approximability of minimizing nonzero variables or unsatisfied relations in linear systems. Theoretical Computer Science 1998;209:237–260.

27. Guyon I, Elisseeff A. An introduction to variable and feature selection. J Mach Learn Res 2003;3:1157–1182.

28. Canter RG, Penney J, Tsai L-H. The road to restoring neural circuits for the treatment of Alzheimer’s disease. Nature 2016;539:187–196.

29. Hyman BT, Phelps CH, Beach TG, et al. National Institute on Aging–Alzheimer’s Association guidelines for the neuropathologic assessment of Alzheimer’s disease. Alzheimer’s & dementia 2012;8:1–13.

30. Corder E, Saunders A, Strittmatter W, et al. Gene dose of apolipoprotein E type 4 allele and the risk of Alzheimer’s disease in late onset families. Science 1993;261:921–923.

31. Morris JC, Roe CM, Xiong C, et al. APOE predicts amyloid-beta but not tau Alzheimer pathology in cognitively normal aging. Annals of neurology 2010;67:122–131.

32. Farrer LA, Cupples LA, Haines JL, et al. Effects of age, sex, and ethnicity on the association between apolipoprotein E genotype and Alzheimer disease: a meta-analysis. Jama 1997;278:1349–1356.

33. Lautrup S, Sinclair DA, Mattson MP, Fang EF. NAD+ in brain aging and neurodegenerative disorders. Cell metabolism 2019;30:630–655.

34. Horgusluoglu E, Nudelman K, Nho K, Saykin AJ. Adult neurogenesis and neurodegenerative diseases: a systems biology perspective. American Journal of Medical Genetics Part B: Neuropsychiatric Genetics 2017;174:93–112.

35. Felsky D, Roostaei T, Nho K, et al. Neuropathological correlates and genetic architecture of microglial activation in elderly human brain. Nature communications 2019;10:1–12.

36. Lee Y-J, Han SB, Nam S-Y, Oh K-W, Hong JT. Inflammation and Alzheimer’s disease. Archives of pharmacal research 2010;33:1539–1556.

37. Mahoney ER, Dumitrescu L, Seto M, et al. Telomere length associations with cognition depend on Alzheimer’s disease biomarkers. Alzheimer’s & Dementia: Translational Research & Clinical Interventions 2019;5:883–890.

38. Wong MW, Braidy N, Poljak A, Pickford R, Thambisetty M, Sachdev PS. Dysregulation of lipids in Alzheimer’s disease and their role as potential biomarkers. Alzheimer’s & Dementia 2017;13:810–827.

39. Suzanne M, Tong M. Brain metabolic dysfunction at the core of Alzheimer’s disease. Biochemical pharmacology 2014;88:548–559.

40. Bourdenx M, Martín-Segura A, Scrivo A, et al. Chaperone-mediated autophagy prevents collapse of the neuronal metastable proteome. Cell 2021;184:2696-2714. e2625.

41. Liu J, Li L. Targeting autophagy for the treatment of Alzheimer’s disease: challenges and opportunities. Frontiers in molecular neuroscience 2019;12:203.

42. Fang EF, Hou Y, Palikaras K, et al. Mitophagy inhibits amyloid-β and tau pathology and reverses cognitive deficits in models of Alzheimer’s disease. Nature neuroscience 2019;22:401–412.

43. Kerr JS, Adriaanse BA, Greig NH, et al. Mitophagy and Alzheimer’s disease: cellular and molecular mechanisms. Trends in neurosciences 2017;40:151–166.

44. Sevigny J, Chiao P, Bussière T, et al. The antibody aducanumab reduces Aβ plaques in Alzheimer’s disease. Nature 2016;537:50–56.

45. Sims R, Hill M, Williams J. The multiplex model of the genetics of Alzheimer’s disease. Nature neuroscience 2020;23:311–322.

46. Schwartzentruber J, Cooper S, Liu JZ, et al. Genome-wide meta-analysis, fine-mapping and integrative prioritization implicate new Alzheimer’s disease risk genes. Nature Genetics 2021;53:392–402.

47. Chia R, Sabir MS, Bandres-Ciga S, et al. Genome sequencing analysis identifies new loci associated with Lewy body dementia and provides insights into its genetic architecture. Nature genetics 2021;53:294–303.

48. Ding Y, Sohn JH, Kawczynski MG, et al. A deep learning model to predict a diagnosis of Alzheimer disease by using 18F-FDG PET of the brain. Radiology 2019;290:456–464.

49. Jo T, Nho K, Saykin AJ. Deep learning in Alzheimer’s disease: diagnostic classification and prognostic prediction using neuroimaging data. Frontiers in aging neuroscience 2019;11:220.

50. Stamate D, Kim M, Proitsi P, et al. A metabolite-based machine learning approach to diagnose Alzheimer-type dementia in blood: Results from the European Medical Information Framework for Alzheimer disease biomarker discovery cohort. Alzheimer’s & Dementia: Translational Research & Clinical Interventions 2019;5:933–938.

51. Bellomo G, Indaco A, Chiasserini D, et al. Machine Learning Driven Profiling of Cerebrospinal Fluid Core Biomarkers in Alzheimer’s Disease and Other Neurological Disorders. Frontiers in neuroscience 2021;15:337.

52. Zhang M, Schmitt-Ulms G, Sato C, et al. Drug repositioning for Alzheimer’s disease based on systematic ‘omics’ data mining. PloS one 2016;11:e0168812.

53. Rodriguez S, Hug C, Todorov P, et al. Machine learning identifies candidates for drug repurposing in Alzheimer’s disease. Nature communications 2021;12:1–13.

54. Veitch DP, Weiner MW, Aisen PS, et al. Understanding disease progression and improving Alzheimer’s disease clinical trials: Recent highlights from the Alzheimer’s Disease Neuroimaging Initiative. Alzheimers Dement 2019;15:106–152.

55. Saykin AJ, Shen L, Yao X, et al. Genetic studies of quantitative MCI and AD phenotypes in ADNI: Progress, opportunities, and plans. Alzheimers Dement 2015;11:792–814.

56. Saykin AJ, Shen L, Yao X, et al. Genetic studies of quantitative MCI and AD phenotypes in ADNI: Progress, opportunities, and plans. Alzheimers Dement 2015;11:792–814.

57. Park YH, Hodges A, Risacher SL, et al. Dysregulated Fc gamma receptor-mediated phagocytosis pathway in Alzheimer’s disease: network-based gene expression analysis. Neurobiol Aging 2020;88:24–32.

58. Horgusluoglu-Moloch E, Nho K, Risacher SL, et al. Targeted neurogenesis pathway-based gene analysis identifies ADORA2A associated with hippocampal volume in mild cognitive impairment and Alzheimer’s disease. Neurobiol Aging 2017;60:92–103.

59. Freedman ML, Reich D, Penney KL, et al. Assessing the impact of population stratification on genetic association studies. Nat Genet 2004;36:388–393.

60. Park YH, Hodges A, Simmons A, et al. Association of blood-based transcriptional risk scores with biomarkers for Alzheimer disease. Neurol Genet 2020;6:e517.

61. Purcell S, Neale B, Todd-Brown K, et al. PLINK: a tool set for whole-genome association and population-based linkage analyses. Am J Hum Genet 2007;81:559–575.

62. Krizhevsky A, Sutskever I, Hinton GE. Imagenet classification with deep convolutional neural networks. Advances in neural information processing systems; 2012: 1097–1105.

63. Hochreiter S, Schmidhuber J. Long short-term memory. Neural computation 1997;9:1735–1780.

64. Zhang J, Li Y, Tian J, Li T. LSTM-CNN Hybrid Model for Text Classification. 2018 IEEE 3rd Advanced Information Technology, Electronic and Automation Control Conference (IAEAC); 2018 12-14 Oct. 2018: 1675–1680.

65. Vaswani A, Shazeer N, Parmar N, et al. Attention is all you need. arXiv preprint arXiv:170603762 2017.

66. Rosenblatt F. The perceptron, a perceiving and recognizing automaton Project Para: Cornell Aeronautical Laboratory, 1957.

67. Rosenblatt F. The perceptron: a probabilistic model for information storage and organization in the brain. Psychological review 1958;65:386.

68. McCulloch WS, Pitts W. A logical calculus of the ideas immanent in nervous activity. The bulletin of mathematical biophysics 1943;5:115–133.

69. Widrow B, Hoff ME. Adaptive switching circuits: Stanford Univ Ca Stanford Electronics Labs, 1960.

70. Minsky M, Papert SA. Perceptrons: An introduction to computational geometry: MIT press, 2017.

71. Werbos PJ. Applications of advances in nonlinear sensitivity analysis. System modeling and optimization: Springer, 1982: 762–770.

72. Werbos PJ. Backwards differentiation in AD and neural nets: Past links and new opportunities. Automatic differentiation: Applications, theory, and implementations 2006:15–34.

73. Rumelhart DE, Hinton GE, Williams RJ. Learning representations by back-propagating errors. nature 1986;323:533–536.

74. LeCun Y, Touresky D, Hinton G, Sejnowski T. A theoretical framework for back-propagation. Proceedings of the 1988 connectionist models summer school; 1988: 21–28.

75. Goodfellow I, Bengio Y, Courville A, Bengio Y. Deep learning: MIT press Cambridge, 2016.

76. Nair V, Hinton GE. Rectified linear units improve restricted boltzmann machines. Icml; 2010.

77. Glorot X, Bordes A, Bengio Y. Deep sparse rectifier neural networks. Proceedings of the fourteenth international conference on artificial intelligence and statistics; 2011: 315–323.

78. Duchi J, Hazan E, Singer Y. Adaptive subgradient methods for online learning and stochastic optimization. Journal of machine learning research 2011;12.

79. Hinton G, Srivastava N, Swersky K. Neural networks for machine learning lecture 6a overview of mini-batch gradient descent. Cited on 2012;14.

80. Kingma DP, Ba J. Adam: A method for stochastic optimization. arXiv preprint arXiv:14126980 2014.

81. Sutskever I, Martens J, Dahl G, Hinton G. On the importance of initialization and momentum in deep learning. International conference on machine learning; 2013: 1139–1147.

82. Breiman L. Random Forests. Machine Learning 2001;45:5–32.

83. Jo T, Cheng J. Improving protein fold recognition by random forest. BMC Bioinformatics 2014;15:S14.

84. Saunders AM, Strittmatter WJ, Schmechel D, et al. Association of apolipoprotein E allele ϵ4 with late-onset familial and sporadic Alzheimer’s disease. Neurology 1993;43:1467–1467.

85. Roses AD, Lutz MW, Amrine-Madsen H, et al. A TOMM40 variable-length polymorphism predicts the age of late-onset Alzheimer’s disease. The Pharmacogenomics Journal 2010;10:375–384.

86. Cervantes S, Samaranch L, Vidal-Taboada JM, et al. Genetic variation in APOE cluster region and Alzheimer’s disease risk. Neurobiology of Aging 2011;32:2107.e2107-2107.e2117.

87. Pruim RJ, Welch RP, Sanna S, et al. LocusZoom: regional visualization of genome-wide association scan results. Bioinformatics 2010;26:2336–2337.

88. Zhao Y, Wang Y, Yang J, et al. Sorting nexin 12 interacts with BACE1 and regulates BACE1-mediated APP processing. Molecular neurodegeneration 2012;7:1–10.

89. Lee J, Retamal C, Cuitiño L, et al. Adaptor protein sorting nexin 17 regulates amyloid precursor protein trafficking and processing in the early endosomes. Journal of Biological Chemistry 2008;283:11501–11508.

90. Gallon M, Clairfeuille T, Steinberg F, et al. A unique PDZ domain and arrestin-like fold interaction reveals mechanistic details of endocytic recycling by SNX27-retromer. Proceedings of the National Academy of Sciences 2014;111:E3604–E3613.

91. Heiseke A, Schöbel S, Lichtenthaler SF, et al. The Novel Sorting Nexin SNX33 Interferes with Cellular PrPSc Formation by Modulation of PrPc Shedding. Traffic 2008;9:1116–1129.

92. Mercado N, Colley T, Baker JR, et al. Bicaudal D1 impairs autophagosome maturation in chronic obstructive pulmonary disease. FASEB BioAdvances 2019;1:688–705.

93. Swan A, Nguyen T, Suter B. Drosophila Lissencephaly-1 functions with Bic-D and dynein in oocyte determination and nuclear positioning. Nature Cell Biology 1999;1:444–449.

